# High and low frequency anterior nucleus of thalamus deep brain stimulation: Impact on memory and mood in five patients with treatment resistant temporal lobe epilepsy

**DOI:** 10.1101/2024.02.14.24302765

**Authors:** Victoria S. Marks, Irena Balzekas, Jessica A. Grimm, Thomas J. Richner, Vladimir Sladky, Filip Mivalt, Nicholas M. Gregg, Brian N. Lundstrom, Kai J. Miller, Boney Joseph, Jamie Van Gompel, Benjamin Brinkmann, Paul Croarkin, Eva C. Alden, Vaclav Kremen, Michal Kucewicz, Gregory A Worrell

**Affiliations:** Department of Neurology, Bioelectronics, Neurophysiology, and Engineering Laboratory, Mayo Clinic, Rochester, MN, United States; Biomedical Engineering and Physiology Graduate Program, Mayo Clinic Graduate School of Biomedical Sciences, Rochester, MN, United States; Mayo Clinic Alix School of Medicine, Rochester, MN, United States; Mayo Clinic Medical Scientist Training Program, Rochester, MN, United States; Faculty of Biomedical Engineering, Czech Technical University in Prague, Prague, Czechia; Faculty of Electrical Engineering and Communication, Department of Biomedical Engineering, Brno University of Technology, Brno, Czechia; Department of Psychiatry and Psychology, Mayo Clinic, Rochester, MN, United States; Department of Neurosurgery, Mayo Clinic, Rochester, MN, United States; Czech Institute of Informatics, Robotics and Cybernetics, Czech Technical University in Prague, Prague, Czechia; Department of Radiology, Mayo Clinic, Rochester, MN, United States; International Clinic Research Center, St. Anne’s University Research Hospital, Brno, Czechia; Department of Biostatistics, Mayo Clinic, Rochester, MN, United States; BioTechMed Center, Brain & Mind Electrophysiology Lab, Multimedia Systems Department, Faculty of Electronics, Telecommunication and Informatics, Gdansk University of Technology, 80-233 Gdansk, Poland

## Abstract

High frequency anterior nucleus of the thalamus deep brain stimulation (ANT DBS) is an established therapy for treatment resistant focal epilepsies. Although high frequency-ANT DBS is well tolerated, patients are rarely seizure free and the efficacy of other DBS parameters and their impact on comorbidities of epilepsy such as depression and memory dysfunction remain unclear. The purpose of this study was to assess the impact of low vs high frequency ANT DBS on verbal memory and self-reported anxiety and depression symptoms. Five patients with treatment resistant temporal lobe epilepsy were implanted with an investigational brain stimulation and sensing device capable of ANT DBS and ambulatory intracranial electroencephalographic (iEEG) monitoring, enabling long-term detection of electrographic seizures. While patients received therapeutic high frequency (100 and 145 Hz continuous and cycling) and low frequency (2 and 7 Hz continuous) stimulation, they completed weekly free recall verbal memory tasks and thrice weekly self-reports of anxiety and depression symptom severity. Mixed effects models were then used to evaluate associations between memory scores, anxiety and depression self-reports, seizure counts, and stimulation frequency. Memory score was significantly associated with stimulation frequency, with higher free recall verbal memory scores during low frequency ANT DBS. Self-reported anxiety and depression symptom severity was not significantly associated with stimulation frequency. These findings suggest the choice of ANT DBS stimulation parameter may impact patients’ cognitive function, independently of its impact on seizure rates.

## Introduction

Epilepsy is associated with a variety of comorbidities including depression, anxiety, and cognitive dysfunction[1–3]. Anterior nucleus of the thalamus deep brain stimulation (ANT DBS) is an established neuromodulatory therapy for treatment resistant epilepsy (TRE) with focal onset seizures[4, 5]. The relationships between different ANT DBS parameters, seizure reductions, and comorbidities such as depression and memory dysfunction remain unclear.

The most established stimulation paradigm for ANT DBS is the one used in the pivotal SANTE trial which involves high frequency, duty cycle stimulation with 145 Hz ON for 1 min and OFF for 5 minutes[5, 6]. Despite initial concerns about subjective worsening of depression and memory symptoms[5], large follow-up studies have largely shown that chronic stimulation with the SANTE paradigm is well tolerated at the group level and associated with improvements in seizure control, executive function, and attention at 7 years following implantation[6, 7]. Since improvements in seizure frequency are associated with long-term ANT DBS, it remains unclear if comorbidity improvements are driven by changes in seizure frequency or are a direct consequence of stimulation.

The stimulation frequency that optimizes seizure control while minimizing side effects for an individual patient remains unknown. DBS has a seemingly paradoxical effect on the brain wherein it can both inhibit local activity and increase efferent output from the stimulated nucleus[8]. Modeling work in DBS for movement disorders indicates that this may be due to a frequency dependent “informational lesion”. During high frequency DBS, the activated output of the nucleus lacks informational content, preventing pathological activity from being transmitted[8]. Why low frequency DBS can also be effective for epilepsy is unclear. The magnitude of the informational lesion may be smaller, or of shorter duration, in low frequency DBS. Stimulation-frequency-specific impacts on comorbidities of epilepsy are unclear as well[9–11]. No significant effect on cognition was found in the SANTE trials[12], but others have found improvement with high frequency ANT stimulation without mention of low-frequency[13].

High frequency ANT DBS has largely been shown to be very well tolerated neuropsychiatrically[6, 14], though adverse effects have been described. Sudden and gradual onset depressive and paranoid symptoms associated with high frequency, duty cycle ANT DBS have been reported, though these symptoms were alleviated by changes in stimulation current or contacts[15, 16]. As alternative stimulation paradigms, such as low frequency ANT DBS are explored, further characterization of parameter-specific neuropsychiatric side effects is needed.

The purpose of this study was to assess the impact of low vs high frequency ANT DBS on free recall verbal memory and self-reported anxiety and depression symptom severity. Five patients with treatment resistant temporal lobe epilepsy were implanted with the investigational Medtronic Summit RC+S™ device which enables ANT DBS and long-term ambulatory intracranial electroencephalographic (iEEG) recordings[17]. The participants were equipped with the Epilepsy Personal Assistant Device (EPAD) system[18], which participants used to complete regular verbal memory tasks and ecological momentary assessments (EMAs) of depression and anxiety symptoms at home. With these data streams, we were able to compare electrographic seizure rates with memory and mood scores collected during high frequency duty cycle and low frequency ANT DBS.

## Methods

### Study design

This research was conducted as part of a study evaluating the safety and feasibility of brain state tracking and modulation in temporal lobe epilepsy (ClinicalTrials.gov Identifier: NCT03946618). This study was approved by the Mayo Clinic Institutional Review Board and the Food and Drug Administration (IDE G180224). Five participants with treatment resistant temporal lobe epilepsy were implanted with the investigational Medtronic Summit RC+S™ DBS system with four leads targeting the bilateral ANT and amygdala-hippocampal formations. The device and integrated cloud-based research environment, called the BrainRISE (Restoration Intelligent Stimulation Ecosystem), for collection of electrophysiology data, patient reports, mood assessments and verbal memory testing using epilepsy patient assistant, a custom software application running on a small handheld computer (EPAD). The EPAD enables bidirectional communication between implanted devices with wireless connectivity and local and cloud computing resource[18, 19]. The EPAD application orchestrates connectivity between multiple wireless capable devices (implant, tablet computer, iPhone, Apple Watch). The BrainRISE platform enabled collection of several continuous and intermittent data streams outlined here, including ambulatory iEEG monitoring, ANT-DBS stimulation parameter changes, seizure detection, IED detection, memory tasks, and ecological momentary assessments. Data were collected from participants for 1-2 years.

### Participants

Study participants all had drug resistant mesial temporal lobe epilepsy involving either the bilateral or dominant hemisphere. Four women (ages 20s-50s) and one man (age 30s) were enrolled. Participant seizure semiology, demographics, and seizure onset zones are available in Table 1.

**Table 1.** Participant demographics. Abbreviations (F: female, M: male, R: right hand dominant, L: left hand dominant, FIAS: focal impaired awareness seizure, GTC: generalized tonic clonic seizure, HC: hippocampus, GAD: generalized anxiety disorder, MDD: major depressive disorder, MDE: major depressive episode). *Patient had a previous left amygdalohippocampectomy and partial temporal lobectomy.

| Participant | Age/ Sex/ Handedness | Baseline seizure frequency | Seizure semiology | Seizure onset zone | Psychiatric comorbidities |
| --- | --- | --- | --- | --- | --- |
| S1 | 50s / F / R+L | 2-3 clusters per week of 5-6/day | FIAS | R HC, L HC, L mid-cingulate | MDD, PD |
| S2 | 20s / F / R | 2-3 per week | FIAS | R HC, L HC remnant* | MDE, GAD |
| S3 | 30s / F / R | Daily | FIAS, GTC | R HC, L HC | MDD |
| S4 | 40s / F / R | 2-3 per week | FIAS | L HC | MDD, GAD |
| S5 | 30s / M / R | 1-2 per month | FIAS, GTC | R HC, L HC | MDD |

### IEEG recording

The system conducted continuous iEEG sampling at 250 or 500 Hz from four bipolar pairs selected from four leads. Data were streamed via Bluetooth to a handheld tablet computer and then to a physician-accessible cloud-based server[19].

### Anterior nucleus of the thalamus deep brain stimulation

Participants received therapeutic ANT DBS within clinically defined parameters. Stimulation frequencies tested included continuous 2, 7, 100, and 145 Hz and duty cycle 145 Hz (1 minute on, 5 minutes off). We grouped 2 Hz and 7 Hz stimulation into the low frequency “lo” condition and 100 Hz, and 145 Hz (both continuous and cycling) in the high frequency “hi” condition. During the memory assessments, all participants began the study on low frequency stimulation that was gradually uptitrated in amplitude (2-6 mA), receiving 4-6 months of low frequency stimulation, and were subsequently transitioned to high frequency duty cycle stimulation for uptitration and stimulation over 4-6 months. Due to variability in patient engagement, impact of COVID-19 pandemic, and availability to participate in behavioral tasks, and data rate, concurrent behavioral measures (Free recall task and ecological momentary assessments) during both high and low frequency stimulation conditions are only available for participants S1 and S4.

### Free recall task

We used a Free Recall Task to assess participants’ verbal memory[20]. The participants completed the tasks on a handheld device in their home environments while iEEG and behavioral data were streamed to the cloud repository. Each participant with lists of 12 randomly chosen nouns, displayed individually. After a distractor task of basic arithmetic, the subject was asked to recall as many of the words as possible. Each session consisted of 15 lists. ANT DBS was paused during the task period. A researcher remained on the phone with the participant for the duration of the task.

### Ecological momentary assessments

One-to-four times a week, participants were prompted on a random day and time to complete the Immediate Mood Scaler 12 (IMS)[21]. The IMS is a 12-item ecological momentary assessment with 7-point Likert scale questions evaluating severity of anxiety and depression symptoms in the moment. Higher total IMS scores indicate less severe symptoms and lower (more negative) scores indicate more severe symptoms. The IMS was presented through the handheld tablet through a licensing agreement with Posit Science (San Francisco, California). The IMS scores were averaged within each week for comparison with the single, weekly, memory score.

### Seizure detection

Hippocampal electrographic seizures were identified in the continuous iEEG time-series by a validated, high-sensitivity automated seizure classifier[17] and visually verified by a board-certified epileptologist (GAW or NMG). The total number of seizures in the calendar week in which the memory task was performed constituted the seizure count for that week.

### Statistical analysis

For overall, group-wise, statistical comparisons, we used a two-sample Wilcoxon Rank Sum test (wilcox.test in R)[22]. We used a linear mixed effect model (LMM) and a generalized linear mixed effect model (GLMM) to investigate the interactions between weekly memory score, seizure count, stimulation frequency, time, and patient. We used an LMM to predict memory, but a GLMM was necessary to predict seizures which are Poisson distributed. Both LMMs and GLMMs are particularly useful when there are mixed effects (i.e. both fixed such as stimulation frequency and random effects such as participants). In all LMMs and GLMMs, time was modeled as a cubic spline, and a random intercept was used for each individual participant. To model memory, we used the *lmer()* function in R to create LMMs with inputs of time, patient, stimulation frequency, memory, and IMS score. To model seizure count, we used the *glmer()* function in R to create Poisson-distributed GLMMS. We tested whether a specific fixed effect had an effect by comparing the likelihood of a model with the predictor of interest in the model to a model without the predictor of interest included. Models were compared using a Likelihood Ratio Test. Because we want to compare models to test a fixed effect, we could not use restricted maximum likelihood estimates (setting *REML=FALSE* for each model), but instead used basic maximum likelihood (ML). All likelihood tests were computed with the ANOVA function with *test = “Chisq”* in R. We completed the analysis on two sets of data. In the first analysis, the input data are limited to weeks where IMS and memory scores were completed in the same week, yielding 100 samples across 5 participants. In the second analysis, IMS data were excluded and every week in which the memory scores were collected was considered, yielding 151 samples across 5 participants. The models tested and their results are included in Tables 2 and 3.

**Table 2.** Model outputs for analysis 1. The results described in this table were derived from weeks in which memory and IMS scores were sampled in the same week (n = 100 across 5 participants). P values for base models comparing the relationship between each variable and time are derived from the fits of the linear mixed effects and generalized linear mixed effects models. P values for comparisons between pairs of models were derived from the Likelihood ratio test (Chi-square). Note, “*”, “**”, and “***” denote p < 0.05, < 0.01, and < 0.001.

| Models | Result | P value |
| --- | --- | --- |
| *IMS as dependent variable* |  |  |
| Question: Is IMS associated with time?<br>IMS ~ time + patient<br>(Second order splines) | ** | 0.0064 |
| Question: Is stimulation associated with IMS when accounting for time?<br>IMS ~ time + patient<br>IMS ~ time + patient + stim | NS | > 0.05 |
| Question: Is memory associated with IMS when accounting for time?<br>IMS ~ time + patient<br>IMS ~ time + patient + memory | NS | > 0.05 |
| Question: Are physician confirmed seizures associated with IMS when accounting for time?<br>IMS ~ time + patient<br>IMS ~ time + patient + seizures | NS | > 0.05 |
| *Memory as dependent variable* |  |  |
| Question: Is memory associated with time?<br>Memory ~ time + patient<br>(First order splines) | ** | 0.0054 |
| Question: Is stimulation associated with memory when accounting for time?<br>Memory ~ time + patient<br>Memory ~ time + patient + stim | *** | 1.3e-06 |
| Question: Is IMS associated with memory when accounting for time?<br>Memory ~ time + patient<br>Memory ~ time + patient + IMS | NS | > 0.05 |
| Question: Are seizures associated with memory when accounting for time?<br>Memory ~ time + patient<br>Memory ~ time + patient + seizures | NS | > 0.05 |
| Question: Do seizures account for the relationship between memory and stimulation?<br>Memory ~ time + patient + stim<br>Memory ~ time + patient + stim + seizures | NS | > 0.05 |
| Question: Does IMS account for the relationship between memory and stimulation?<br>Memory ~ time + patient + stim<br>Memory ~ time + patient + stim + IMS | NS | > 0.05 |
| *Seizures as dependent variable* |  |  |
| Question: Are seizures associate with time?<br>Seizures ~ time + patient<br>(Not significant through 6<sup>th</sup> order splines) | NS | > 0.05 |
| Question = Are seizures associated with stimulation when accounting for time?<br>Seizures ~ time + patient<br>Seizures ~ time + patient + stim | NS | > 0.05 |
| Question = Are seizures associated with memory when accounting for time?<br>Seizures ~ time + patient<br>Seizures ~ time + patient + memory | NS | > 0.05 |
| Question = Are seizures associated with IMS when accounting for time?<br>Seizures ~ time + patient<br>Seizures ~ time + patient + IMS | NS | > 0.05 |

**Table 3.** Model outputs for analysis 2. The results described in this table were derived from all the weeks in which memory was sampled (n = 151 across 5 participants). P values for base models comparing the relationship between each variable and time are derived from the fits of the linear mixed effects and generalized linear mixed effects models. P values for comparisons between pairs of models were derived from the Likelihood ratio test (Chi-square). Note, “*”, “**”, and “***” denote p < 0.05, < 0.01, and < 0.001.

| Models | Result | P value |
| --- | --- | --- |
| *Memory as dependent variable* |  |  |
| Question: Is memory associated with time?<br>Memory ~ time + patient<br>(First order splines) | ** | 0.0020 |
| Question: Is stimulation associated with memory when accounting for time?<br>Memory ~ time + patient<br>Memory ~ time + patient + stim | *** | 5.1e-09 |
| Question: Are seizures associated with memory when accounting for time?<br>Memory ~ time + patient<br>Memory ~ time + patient + seizures | NS | > 0.05 |
| Question: Do seizures account for the relationship between memory and stim?<br>Memory ~ time + patient + stim<br>Memory ~ time + patient + stim + seizures | NS | > 0.05 |
| *Seizures as dependent variable* |  |  |
| Question: Are seizures associated with time?<br>Seizures ~ time + patient | ** | 0.0043 |
| Question = Are seizures associated with stimulation when accounting for time?<br>Seizures ~ time + patient<br>Seizures ~ time + patient + stim | NS | > 0.05 |
| Question = Are seizures associated with memory when accounting for time<br>Seizures ~ time + patient<br>Seizures ~ time + patient + memory | NS | > 0.05 |

### Data availability statement

All data produced in the present study are available upon reasonable request to the authors and data sharing agreement to protect patient privacy.

## Results

The study design is summarized in Figure 1a. Participants received the 4-lead DBS implant. Following an approximate one-month period without stimulation, therapeutic ANT DBS was initiated, and participants received both high and low ANT DBS during the study. Concurrently, participants completed the free recall verbal memory task once weekly and the IMS 1-4 times weekly. Engagement with the memory task and self-reported anxiety and depression symptoms varied between participants. Four of the five participants underwent both low- and high frequency ANT-DBS and the number of weeks where participants completed both the memory task and mood scores under each stimulation paradigm varied. Memory scores samples captured 151 weeks across the five participants, but for only 100 of those weeks were concurrent IMS scores available. Therefore, we separated the analysis into two parts. In the first analysis, we used 100 samples from the 5 participants, accommodating both IMS and memory scores as variables. In the second analysis, we used all 151 memory scores, but excluded IMS as a variable. Figure 2a shows the number of behavioral samples (weeks where participants completed both the memory task and mood scores in the same week) per participant per stimulation frequency. Only participants S1 and S4 completed all behavioral assessments during both high and low frequency ANT DBS.

**Figure 1.**
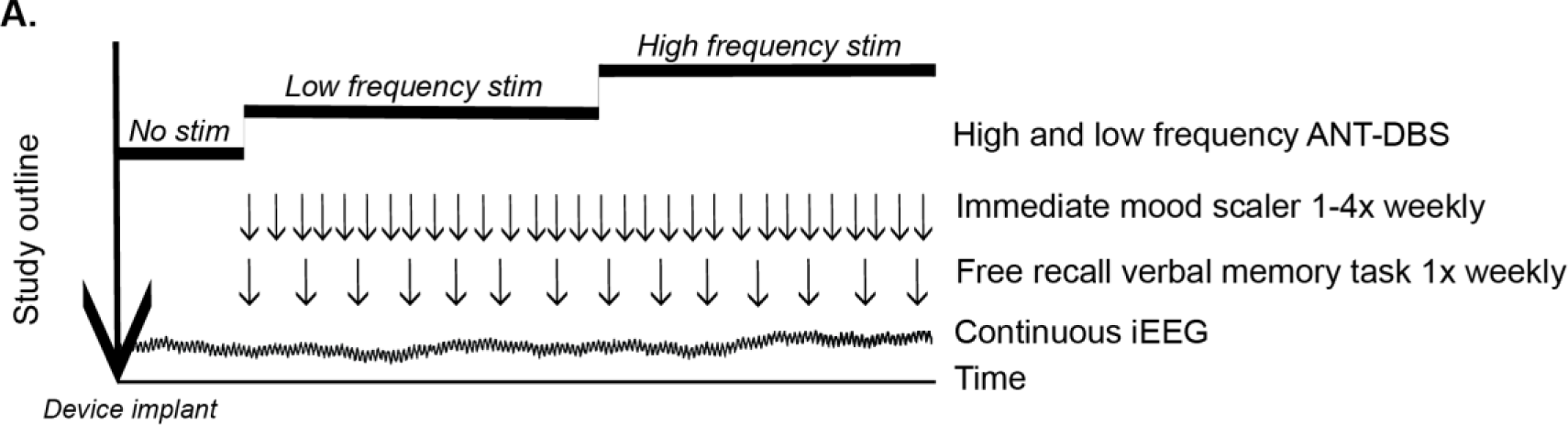
Study schematic and summary. A) Study outline describing timeline from device implant and concurrent data streams collected during the study period. The periods of high and low frequency ANT DBS are depicted as approximations of the stimulation periods which varied between participants. Within each stimulation frequency, the stimulation amplitude was gradually uptitrated week by week based on patient-specific side effects. Consistent behavioral samples are depicted in the figure, though the number of consecutive weeks in which both the verbal memory task and immediate mood scaler were completed varied between participants.

**Figure 2.**
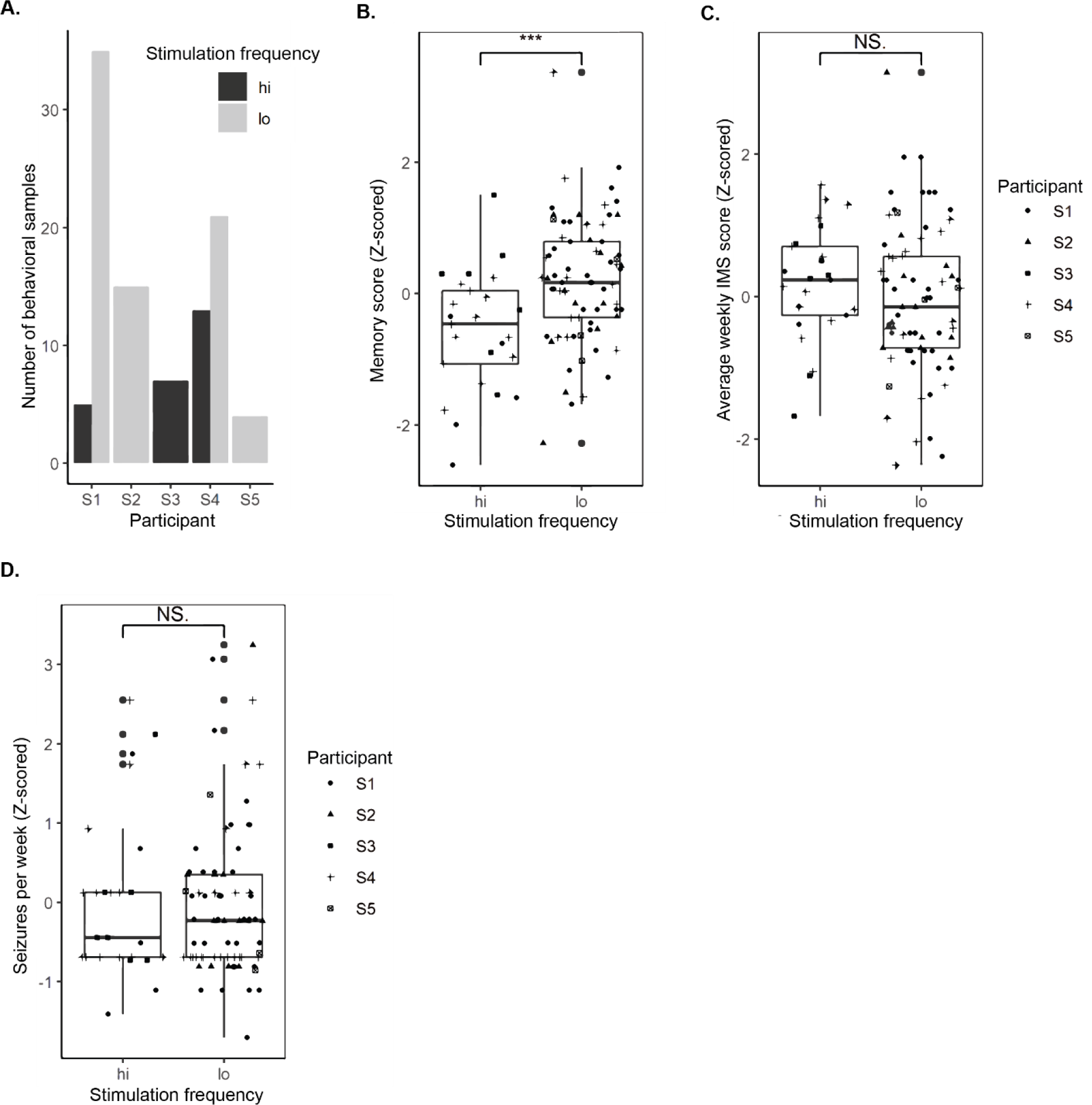
A) Bar plots describing the number of weeks where each participant completed both the memory task and IMS, per ANT DBS stimulation frequency. “Hi” denotes 100 and 145 Hz duty cycle stimulation. “Lo” denotes 2 and 7 Hz continuous stimulation. B) Memory score comparison across all subjects during high and low frequency stimulation. C) IMS score comparison across all subjects during high and low frequency stimulation. D) Number of seizures per week during high and low frequency stimulation. Note, “*”, “**”, and “***” denote p < 0.05, < 0.01, and < 0.001.

In group-wise comparisons, free recall verbal memory scores were significantly higher during low frequency stimulation than during high frequency stimulation (p < 0.001) (Figure 2b). Of note, stimulation was paused for the duration of the task, so participants were not actively receiving stimulation at that time. The average weekly IMS score did not differ significantly between high and low frequency stimulation (p > 0.05) (Figure 2c). Additionally, weekly seizure count for the periods sampled in the study did not differ significantly between the high and low frequency stimulation periods (Figure 2d).

To further investigate the impact of stimulation frequency on memory, we performed further evaluation with a mixed effects model to accommodate time as a variable. Seizures in epilepsy show multiday periodicities[23], making it unlikely that weekly seizure counts are independent, identically distributed variables. We used the linear mixed effect models to investigate the interactions between time, memory score, IMS score, seizure count, stimulation frequency, time, and patient factors.

In the first analysis, we included time, patient factors, memory score, IMS score, stimulation frequency, and seizures as variables. This analysis only included weeks where both memory and IMS scores were collected (100 samples). The results are described in Table 2. There was no significant association between IMS score and stimulation frequency, memory score, or seizures.

Stimulation frequency was significantly associated with memory score (p < 0.001) with higher memory scores during low frequency stimulation. Memory and seizures were not significantly associated, neither were stimulation frequency and seizures. There was no significant difference between the model relating memory to stimulation and the model relating memory to stimulation and seizures, indicating that the relationship between memory and stimulation was not driven by seizures. Altogether, memory score was significantly associated with stimulation frequency, with improved memory performance at low frequency stimulation.

In the second analysis we included time, patient factors, memory score, stimulation frequency, and seizures. All weeks in which the memory score was collected were included. The results are described in Table 3. Stimulation frequency was significantly associated with memory (p < 0.001). Stimulation frequency was not significantly associated with seizures. There was no significant difference between the model relating memory to stimulation and the model relating memory to stimulation and seizures, indicating that the relationship between memory and stimulation was not driven by seizures. Both analyses support the finding that memory score and stimulation frequency are significantly associated, independently of seizures, with better memory performance and low frequency stimulation.

## Discussion

We compared weekly free recall verbal memory scores, self-reported depression and anxiety symptoms, and seizure counts in five patients undergoing long-term high frequency and low frequency ANT DBS for drug resistant mesial temporal lobe epilepsy. Memory performance was significantly better during low frequency than high frequency ANT DBS. Self-reported depression and anxiety symptoms were not significantly associated with stimulation frequency. This finding indicates that the choice of DBS stimulation parameter may impact patients’ cognitive function, independently of depression and anxiety symptoms and of its impact on seizure rates.

The ANT is an important node in the limbic system which is involved in both memory formation, mood, and propagation of epileptiform activity[24–27]. Long-term, high-frequency ANT-DBS has previously been associated with subjective declines in memory performance, but not with declines in objective neuropsychological measures[12]. In fact, at 7 year follow-up as part of the SANTE study, patients have been shown to have improved executive function and attention[12]. Although we are unable to similarly compare memory scores to pre-implant baselines in our analysis of the Free Recall task, our findings support the notion that different stimulation frequencies may modulate limbic circuitry and memory function differently. It is possible that in general terms, ANT-DBS and the associated reduction in seizures is cognitively advantageous, yet when comparing specific stimulation frequencies, low-frequency stimulation may have less of an impact on memory function.

Our findings build upon a preliminary verbal memory analysis of a subset of this patient cohort that applied a different model to patients individually[28]. In this publication by Marks and colleagues, a GLM was used to make one-week-ahead prediction of memory score based on the current and two prior week’s seizure counts and current week’s memory score[28]. Using this approach, the model predictions were strongly correlated with free-recall memory outcomes for only one participant[28]. When considered overall across the entire study period, memory score and weekly seizure rate were not significantly correlated[28]. Our analysis took further steps to incorporate all participants with patient-specific intercepts and accommodate both seizure burden and stimulation frequency as possible contributing or confounding factors. Of the variables tested, we found that stimulation frequency (low vs high frequency ANT DBS) was the main driver of memory performance. Although patients with epilepsy have been shown to have worse memory function such as accelerated long-term forgetting[29], our findings that seizure counts and free recall verbal memory scores on a weekly timescale are not significantly correlated indicate that the impact of seizures on memory may depend on the timescale at which they are assessed, the specific type of memory processing evaluated, be it formation or consolidation, and concurrent therapy be it stimulation or medication.

In this study, stimulation parameter selection was largely driven by clinical assessment of patient response to stimulation. Future studies would benefit from testing a wider range of stimulation frequencies and paradigms and re-crossover elements to determine if direction of stimulation change drives the effect on memory performance (e.g., will patients perform better on memory tasks during high frequency stimulation if they have first received low frequency stimulation) due to long-term changes in plasticity from chronic stimulation.

A key limitation of this study was the absence of a no-stimulation control for the free recall memory task and IMS assessments. Unfortunately, these features were not initially available within the BrainRISE platform at the time of participant enrollments. Our findings suggest that when comparing ANT-DBS frequencies, free recall memory performance is better during low-frequency than during high-frequency stimulation, and that this relationship is independent of seizures. Yet, we were unable accommodate memory scores and seizure rates during periods where patients were not receiving ANT-DBS, prior to DBS surgery, for example. It is possible that ANT-DBS reduced seizure frequency compared to baseline similarly enough that more subtle associations between seizures and memory were obscured. Regardless, the potential impact of different DBS parameters on important cognitive processes is worthy of further consideration.

Additional limitations include the small cohort size and limited stimulation frequency exposure across participants. The lack of significant association between seizures and stimulation frequency may be attributable to the small cohort, variable duration of data collection in each participant, and variable seizure counts between participants ranging from zero to as many as twenty-two seizures in one week. Focal seizures have been previously shown to have multiday cycles with periods on the order of weeks and months[23]. Multiday cycles have been described previously in one participant from this cohort[30]. The phase of the seizure cycle in which a stimulation parameter and memory were assessed likely influenced the observed seizure count, independently of the stimulation frequency. Both low[31, 32] and high frequency[6] brain stimulation have been shown to reduce seizure activity, but a long-term, randomized control trial directly comparing each paradigm will be necessary to clarify these relationships.

Our decision to average IMS scores within each week may have obscured some variability in the IMS signal. As such, our negative findings regarding IMS score, memory, stimulation, and seizures should be interpreted cautiously. The literature comparing self-reported anxiety and depression symptoms with seizures and DBS stimulation has varied findings depending on the psychological measure, sampling frequency, method of seizure quantification be it patient self-reports or electrographic seizure detections, and study duration be it short term DBS parameter changes or long term DBS therapy[33]. Further evaluation with denser sampling of self-reported psychiatric symptoms with additional quantitative metrics such as actigraphy or electrodermal activity may be necessary to better evaluate psychiatric symptomatology in patients receiving chronic ANT-DBS.

In conclusion, this study is the first to densely track markers of comorbidities of epilepsy including cognitive function and self-reported depression and anxiety symptoms during chronic stimulation therapy with concurrent local field potential recordings. Our findings support the concept that the choice of stimulation frequency likely impacts brain processes beyond absolute seizure counts. Most importantly, this study highlights the value in holistic and comprehensive approaches to research in epilepsy treatment, emphasizing both seizure frequency and neuro-cognitive processes that impact quality of life.

